# Genome-wide association study identifies *APOE* and *ZMIZ1* variants as mitophagy modifiers in Lewy body disease

**DOI:** 10.1101/2023.10.16.23297100

**Authors:** Xu Hou, Michael G. Heckman, Fabienne C. Fiesel, Shunsuke Koga, Alexandra I. Soto-Beasley, Jens O. Watzlawik, Jing Zhao, Rebecca R. Valentino, Patrick W. Johnson, Launia J. White, Zachary S. Quicksall, Joseph S. Reddy, Jose Bras, Rita Guerreiro, Na Zhao, Guojun Bu, Dennis W. Dickson, Owen A. Ross, Wolfdieter Springer

## Abstract

The PINK1-PRKN pathway mediates a critical quality control to maintain mitochondrial health and function. Together the kinase-ligase pair identifies and decorate damaged mitochondria with phosphorylated ubiquitin (p-S65-Ub). This selective label serves as the mitophagy tag and facilitates their degradation via autophagy-lysosome system. While complete loss of PINK1 or PRKN function causes early-onset Parkinson disease, much broader mitophagy impairments are emerging across neurodegenerative disorders. We previously found age- and disease-dependent accumulation of p-S65-Ub signal in the hippocampus of autopsy brains with Lewy body disease (LBD). However, the contribution of genetic variation to mitochondrial damage and p-S65-Ub levels remains unknown in LBD cases. To identify novel regulators of PINK1-PRKN mitophagy in LBD, we performed an unbiased genome-wide association study of hippocampal p-S65-Ub level with 1,012 autopsy confirmed LBD samples. Using an established, mostly automated workflow, hippocampal sections were immunostained for p-S65-Ub, scanned, and quantified with unbiased algorithms. Functional validation of the significant hit was performed in animal model and human induced pluripotent stem cells (hiPSCs).

We identified a strong association with p-S65-Ub for *APOE4* (rs429358; *β*: 0.50, 95% CI: 0.41 to 0.69; *p*=8.67x10^-25^) and a genome-wide significant association for *ZMIZ1* (rs6480922; *β*: -0.33, 95% CI: -0.45 to -0.22; *p*=1.42x10^-8^). The increased p-S65-Ub levels in *APOE4*-carrier may be mediated by both co-pathology-dependent and -independent mechanisms, which was confirmed in Apoe-targeted replacement mice and hiPSC-derived astrocytes. Intriguingly, *ZMIZ1* rs6480922 also significantly associated with increased brain weight and reduced neuropathological burden indicating a potential role as a resilience factor. Our findings nominate novel mitophagy regulators in LBD brain (*ZMIZ1* locus) and highlight a strong association of *APOE4* with mitophagy alteration. With *APOE4* being the strongest known risk factor for clinical Alzheimer’s disease and dementia with Lewy bodies, our findings suggest a common mechanistic link underscoring the importance of mitochondrial quality control.

## Introduction

Parkinson’s disease (PD), PD with dementia (PDD), and dementia with Lewy bodies (DLB) form a spectrum of neurodegenerative diseases with heterogenous clinical presentation, including parkinsonism, progressive cognitive decline/dementia, and other non-motor symptoms.^1^ While some controversy persists in their distinction, all three clinical disorders present with the abnormal accumulation of alpha-synuclein (αSyn) in the form of Lewy bodies (LBs) and Lewy neurites in the brain, and are thus together pathologically defined as Lewy body disease (LBD).^1^ Many LBD patients additionally exhibit varying degrees of Alzheimer’s disease (AD) co-pathology including amyloid β (Aβ) senile plaques (SP) and neurofibrillary tangles (NFT) formed by hyperphosphorylated tau (p-tau).^1^ More than six million individuals are affected by LBD globally, and there is no cure or effective therapy to slow disease progression as current treatments only relieve symptoms.^2^

To better understand disease etiology and explore new clinical biomarkers and drug targets, efforts from multiple genome-wide association studies (GWAS) have consistently demonstrated a significant genetic contribution to the development of PD and DLB.^3–5^ These clinical conditions share overlapping common genetic risk factors, including variation in the *SNCA* and *GBA* genes, but also have distinct genetic components such as the *APOE4*, which is a strong risk factor for DLB but not PD, and the MAPT *H1* haplotype, a strong risk factor for PD but not DLB.^3–5^ However, although genetic susceptibility factors for clinical LB disorders have been rigorously studied, genetic modifiers of neuropathological features in LBD are not well understood. The most prominently affected biological pathways that contribute to LBD include alterations in lipid homeostasis, mitochondrial dysfunctions, and lysosomal impairments.^6^

The ubiquitin (Ub) kinase-ligase pair PINK1-PRKN directs a cytoprotective quality control pathway by selectively removing defective mitochondria to maintain a healthy and functional mitochondrial pool.^7^ Upon stress, PINK1-PRKN label terminally damaged mitochondria with phosphorylated Ub (p-S65-Ub), which leads to their elimination via the autophagy-lysosome system. Complete loss of *PINK1* or *PRKN* gene function abrogates activation of mitophagy and causes autosomal recessive early-onset PD.^7^ However, mitophagy flux can also be stalled in lysosomes resulting in the abnormal build-up of p-S65-Ub labeled, dysfunctional mitochondria. In autopsy brain, the mitophagy tag increases with normal aging and independently with LBD or AD in which p-S65-Ub levels were strongly associated particularly with the early stages of the respective neuropathologies.^8–10^ Given that impaired mitophagy might contribute to LB formation and neurodegeneration,^6^ p-S65-Ub level may not only serve as a marker for the dynamic PINK1-PRKN pathway but also as a novel, quantitative trait in LBD.

Currently, it remains unknown as to how genetic variation may influence levels of p-S65-Ub in LBD cases. To dissect the underlying genetic architecture linked to mitophagy alterations in LBD, herein we performed a large-scale two-stage GWAS of hippocampal p-S65-Ub levels in 1,012 pathologically confirmed autopsy LBD brains (**Fig. 1**) and identified significant associations for both *APOE* rs429358 (*APOE4*) and *ZMIZ1* rs6480922. We then evaluated the potential neuropathological changes associated with the identified variants, and functionally validated pathology-independent *APOE4* effects on mitophagy in mouse and induced pluripotent stem cell (iPSC) models. Together, our study provides important new insight into LBD pathogenesis and nominates potential therapeutic targets for early disease management.

**Figure 1.**
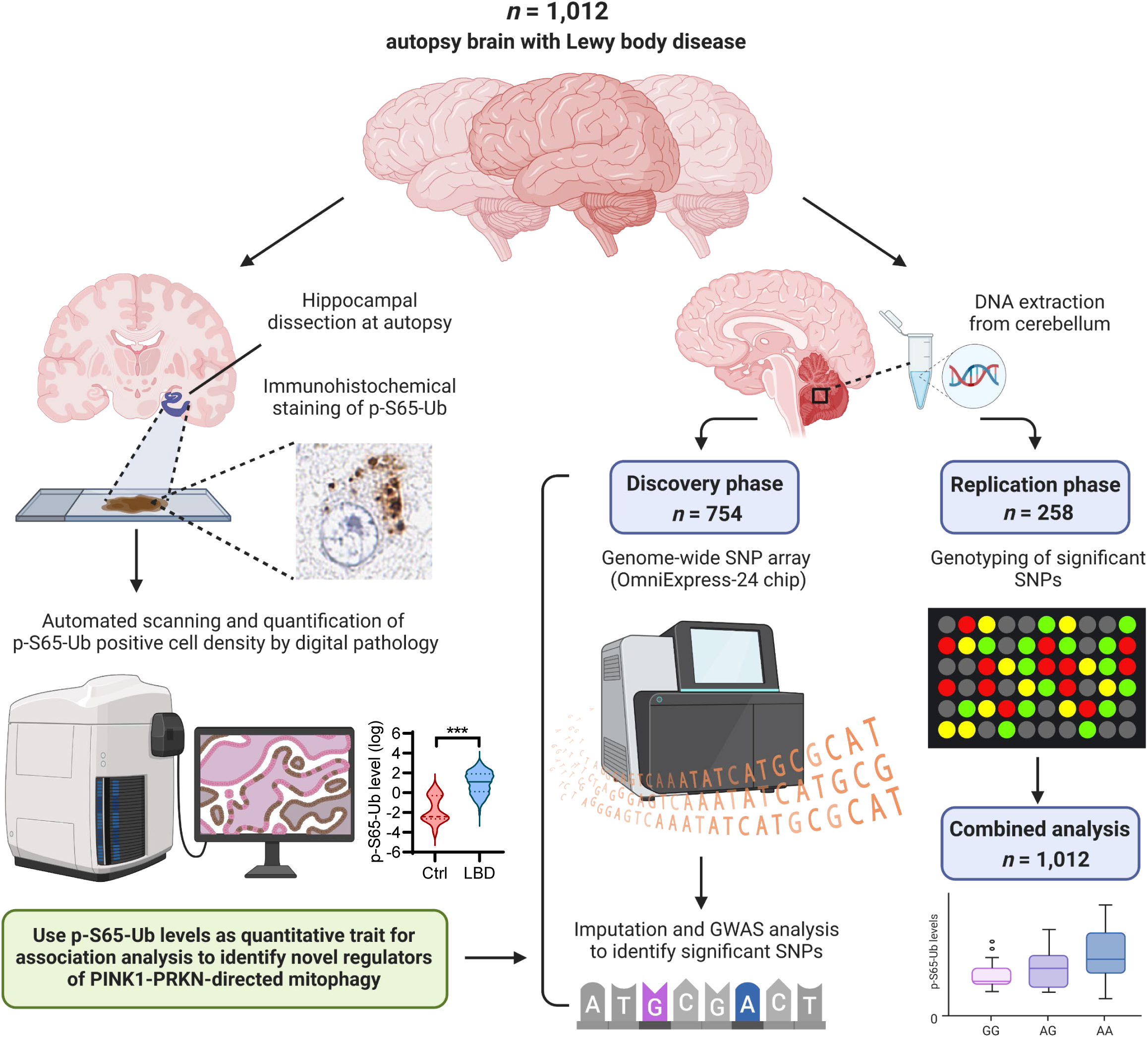
Workflow of the two-step GWAS analysis using hippocampal p-S65-Ub levels as quantitative trait. A total of 1,021 autophagy brain with pathological confirmed LBD were obtained from the brain bank. Hippocampal region was dissected from the fixed hemibrain during autopsy and immunostained for p-S65-Ub by autostainer. Stained hippocampal sections were then scanned and p-S65-Ub positive cell density were quantified using unbiased digital pathology. Representative images of p-S65-Ub positive cells and levels of hippocampal p-S65-Ub cell density in control and LBD cases were shown. DNA was extracted from frozen cerebellar tissue from the same cohort. In the discovery phase, genome-wide SNP array was performed in 754 LBD cases. p-S65-Ub levels were used a quantitative trait for genome-wide association analysis to identify regulators of PINK1-PRKN-directed mitophagy. In the replication phase, significant SNPs were genotyped in the rest 258 LBD cases. Finally, a combined analysis was performed in the entire cohort. This figure is created with BioRender.com.

## Materials and Methods

### Study design and subjects

A total of 1,012 LBD autopsy brains that were obtained from the Mayo Clinic Florida brain bank between 1998 and 2021 were included and divided into discovery series (*n*=754) and independent replication series (*n*=258). All cases were unrelated and self-reported non-Hispanic Caucasians. All brains were examined in a systematic and standardized manner by a single neuropathologist (DWD). Available demographic and neuropathological information included age at death, sex, brain weight, LBD subtype (brainstem, transitional, or diffuse), Braak NFT stage (0-VI), and Thal amyloid phase (0-5), as well as αSyn burden (by immunohistochemistry) and densities of SPs and NFTs (by thioflavin S) in the hippocampus (**Supplemental Table 1-2)**. All brain samples are from autopsies performed after approval by the legal next-of-kin. Research on de-identified postmortem brain tissue is considered exempt from human subjects’ regulations by the Mayo Clinic Institutional Review Board.

### Genome-wide association and replication

Frozen cerebellum brain tissue was collected from all cases and genomic DNA was extracted using Autogen Flex Star (Holliston, MA) methods. LBD cases from discovery series were genotyped in the Illumina OmniExpress-24 genotyping array (Illumina, San Diego, CA, USA). Subject-level exclusion criteria based on GWAS data included a call rate <98%, significant relatedness (PI_HAT>0.25), and non-Caucasian ancestry as determined by ADMIXTURE.^11^ Variants were excluded if they had a call rate <98%, a Hardy-Weinberg equilibrium (HWE) *p*<1x10^-5^, or a minor allele frequency (MAF) <1%. Genome-wide imputation was performed with the TOPMed Imputation Server.^12^ Following imputation, the aforementioned variant-level exclusion criteria were utilized, with the addition of excluding variants with an imputation r^2^ <0.8; this resulted in a total of 8,696,291 variants available for analysis. In the replication series, all samples were genotyped using a custom-designed MassARRAY^®^ System iPlex assay and 2 custom Taqman SNP genotyping assays (rs429358 and rs76354500). All genotype call rates were >99% and there was no evidence of a departure from Hardy-Weinberg equilibrium (all *p*≥0.10).

### Statistical analysis of GWAS data

For the discovery and the replication series, single-variant associations with p-S65-Ub level were examined using linear regression models that were adjusted for age at death, sex, hippocampal region, and the top five principal components (PCs) of genetic data (the top five PCs were not adjusted for in the replication series). p-S65-Ub level was assessed on the natural logarithm scale owing to its skewed distribution. Variants were examined under an additive model (i.e., effect of each additional minor allele). Regression coefficients (denoted as *β*) and 95% confidence intervals (CIs) were estimated and are interpreted as the change in p-S65-Ub level (on the natural logarithm scale) per each additional minor allele of the given variant. For variants that were included in both the discovery series and the replication series, the results of association analyses were combined to obtain regression coefficients, 95% CIs, and p-values using a random-effects meta-analysis with inverse-variance weighting. The results of this meta-analysis combining the two series were considered to be the primary analysis, given that joint analysis of GWAS’s with a discovery/replication approach has been shown to be more efficient than focusing on the replication series alone; (see **Fig. 1** for an overview of the workflow).^13^

In the discovery series and in the combined-analysis, *p*<5x10^-8^ were considered as genome-wide significant, while *p*<1x10^-6^ were considered as displaying suggestive evidence of an association. All statistical tests were two-sided. Statistical analyses were performed using PLINK version 1.9 and R Statistical Software (version 4.1.2).

### Immunohistochemistry and image analysis

Immunohistochemical staining of paraffin embedded postmortem brain tissue containing anterior or posterior hippocampus was performed as previously described (see Supplemental Methods).^8^ After staining, all sections were scanned with an Aperio AT2 digital pathology scanner (Leica Biosystems, Wetzlar, Germany) and then traced and quantified using optimized Aperio algorithms to count the positive cell number followed by manual quality control.

### Apoe-targeted replacement mice and Meso Scale Discovery electrochemiluminescence assays (MSD)

Apoe-targeted replacement (Apoe-TR) mice in which murine *Apoe* gene locus is replaced with human *APOE* gene (either E3 or E4 allele) were purchased from Taconic and used here.^14^ All animal procedures were approved by the Mayo Clinic Institutional Animal Care and Use Committee (IACUC) in accordance with the National Institutes of Health. Age- and sex-matched Apoe-TR mice were anesthetized at 3 or 22 months of age (*n*=7-8 mice/genotype/age group). Brains were quickly removed after transcardial perfusion with phosphate-buffered saline (PBS, pH 7.4), divided along the sagittal plane, and then snap-frozen in liquid nitrogen. For protein extraction, hemi-brain from Apoe-TR mice were homogenized in RIPA buffer (50 mM Tris, pH 8.0, 150 mM NaCl, 0.1% SDS, 0.5% deoxycholate, 1% NP-40). After 30 min incubation on ice, samples were centrifuged for 15 min at 14,000 rpm at 4°, and the supernatant was collected as total brain lysate. Protein content was determined by BCA assay (Thermo Fisher, 23225). p-S65-Ub levels were measured by MSD as previous described (see Supplemental Methods) and read on the MESO QuickPlex SQ 120 (Meso Scale Diagnostics, Rockville, MD, USA).^15^

### Differentiation of human iPSCs into astrocytes and western blot

Human iPSCs from normal individuals with homozygous *APOE3* or *APOE4* genotype were generated under approved IRB protocols with consent for research purpose.^16^. The generation of neural progenitor cells (NPCs) and the differentiation into astrocyte were performed as previously described (see Supplemental Materials).^16^ At day 30, the NPC-derived astrocytes were treated with 10 μM carbonyl cyanide m-chlorophenyl hydrazone (CCCP; Sigma Aldrich, C2759) for 0 or 8 h and then harvested for western blot analyses (see Supplemental Methods) to probe with primary antibodies against p-S65-Ub (in-house, 1:10,000)^17^, PINK1 (Cell Signaling Technology, 6946; 1:1,000), and GAPDH (Meridian Life science, H86504M; 1:150,000).

### Other statistical analysis

Associations of *APOE* rs429358 (*APOE4*) and *ZMIZ1* rs6480922 with brain weight, αSyn burden, and densities of SPs and NFTs were examined using linear regression models with adjustment for age at death and sex. αSyn burden was evaluated on the natural logarithm scale while SP and NFT density were examined on the cube root scale owing to the skewed distributions of these measures. β coefficients and 95% CIs were estimated and are interpreted as previously described. For biochemical measures in animals and cells, unpaired t-tests were used for comparison between *APOE3* and *APOE4* groups, and *p*<0.05 were considered as statistically significant. All statistical tests were two-sided. Statistical analyses were performed using R Statistical Software (version 4.1.2) and GraphPad Prism (GraphPad Software; version 9).

## Results

### GWAS identified variants associated with hippocampal p-S65-Ub levels in LBD brains

In our GWAS analysis of p-S65-Ub levels involving 8,696,291 variants and where all regression models were adjusted for age at death, sex, hippocampal region, and the top 5 PCs, we observed a λ value equal to 1.020, indicating good control of population stratification. A Manhattan plot is displayed in **Fig. 2A**. We identified one genome-wide significant association with p-S65-Ub level, *APOE* rs429358 (*β*: 0.50, 95% CI: 0.39 to 0.62; *p*=1.18x10^-17^), and an additional six suggestive associations (**Table 1**). Reanalysis of the GWAS in non-*APOE4* carriers only (*n*=373) identified another suggestive association for *TMEM123* rs13377311 (*β*: 0.73, 95% CI: 0.46 to 1.00; *p*=1.28x10^-7^; **Table 1** and **Supplemental Fig. 1**). These eight genome-wide significant or suggestive variants were then assessed in the replication series, where there was again a strong association between *APOE* rs429358 and p-S65-Ub level (*β*: 0.52, 95% CI: 0.32 to 0.72; *p*=9.92x10^-7^; **Table 1**). Some evidence of an independent replication was also observed for *ZMIZ1* rs6480922 where the association was almost nominally significant (*β*: -0.23, 95% CI: -0.48 to 0.01; *p*=0.065; **Table 1**). Genotype, allele counts, and frequencies of variants identified and assessed in the discovery and replication series are shown in **Supplemental Table 3a and 3b**, respectively.

**Figure 2.**
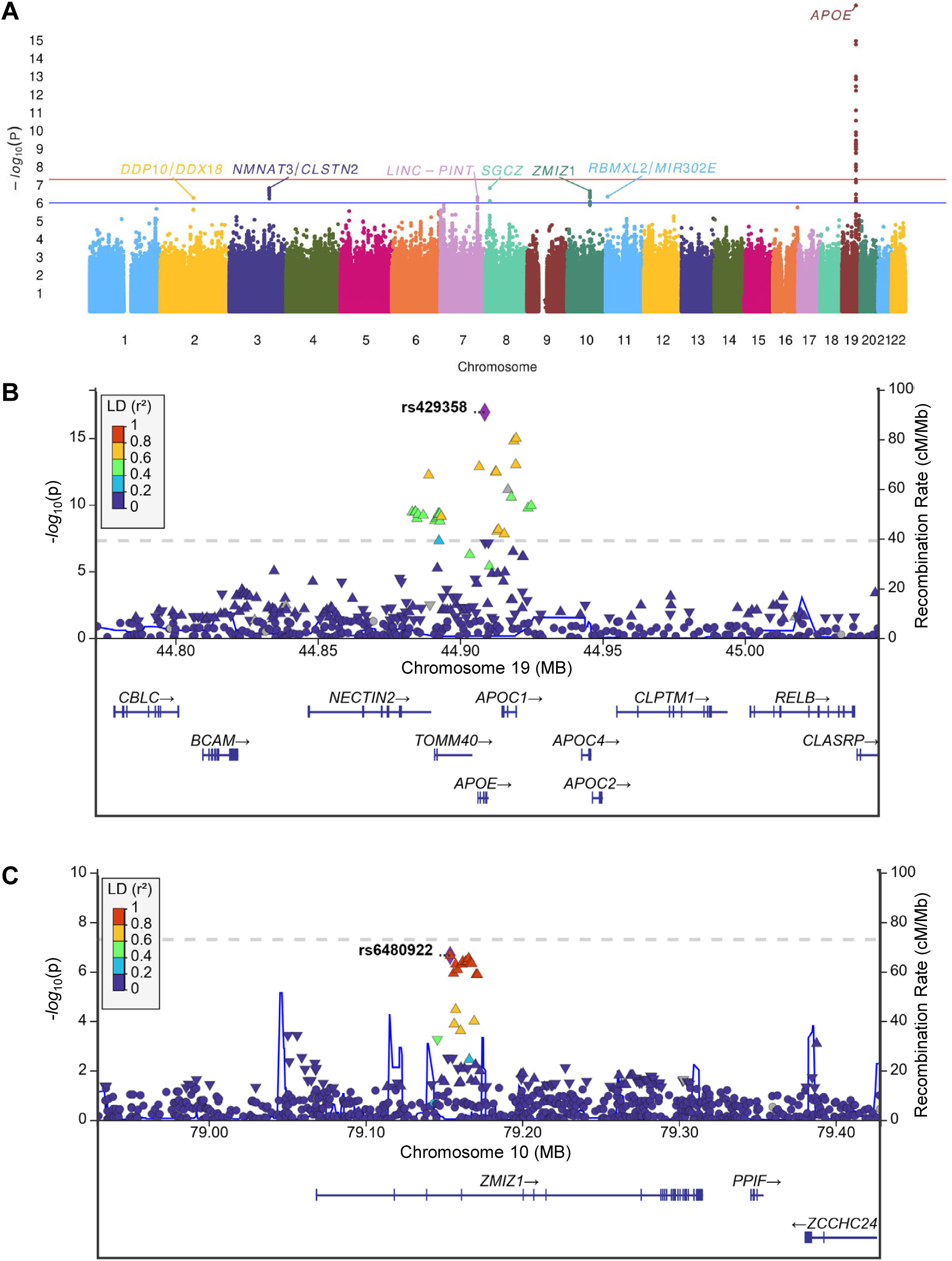
GWAS analysis identified two distinct mitophagy modifiers in LBD autopsy brain. **(A)** Manhattan plot showing genome-wide p-values of association with p-S65-Ub levels. The Y axis shows -log10 p-values of 8,696,291 SNPs and the X axis shows their chromosomal positions. The red solid line indicates the p-value threshold used to define a genome-wide significant association (*p*<5x10^-8^), and the blue solid line indicates the p-value threshold used to define a “suggestive” association (*p*<1x10^-6^). (**B, C**) Locus Zoom plot showing association of variants at the *APOE* (B) and *ZMIZ1* (C) loci with p-S65-Ub levels. The most significant variants (rs429358 and rs6480922) are indicated in purple. The association p-value is shown on the Y-axis and linear position on the chromosome on the X axis. Each point on the plot represents one variant; the colors of the points indicate the linkage disequilibrium (r^2^) value with the index variant (rs429358 or rs6480922). The direction of the point is the direction of *β* for all variants that have a nominal p-value. Points that are missing linkage disequilibrium information are shown in grey.

**Table 1.** Genome-wide or suggestive associations with p-S65-Ub level.

|  |  |  |  | Discovery series (n=754) |  |  |  | Replication series (n=258) |  |  | Combined analysis (n=1,012) |  |
| --- | --- | --- | --- | --- | --- | --- | --- | --- | --- | --- | --- | --- |
| Chr. | Position | Variant | Gene or closest gene<sup>a</sup> | MA | MAF | β (95% CI) | p-value | MAF | β (95% CI) | p-value | β (95% CI) | p-value |
| Genome-wide significant associations in discovery series (p<5 × 10<sup>-8</sup>) |  |  |  |  |  |  |  |  |  |  |  |  |
| 19 | 44908684 | rs429358 | APOE | C | 30.0% | 0.50 (0.39, 0.62) | 1.18 × 10<sup>-17</sup> | 29.1% | 0.52 (0.32, 0.72) | 9.92 × 10<sup>-7</sup> | 0.50 (0.41, 0.60) | 8.67 × 10<sup>-25</sup> |
| Suggestive associations in discovery series (p<1 × 10<sup>-6</sup>) |  |  |  |  |  |  |  |  |  |  |  |  |
| 2 | 116947120 | rs6712544 | DPP10 & DDX18<sup>a</sup> | A | 4.8% | -0.64 (-0.89, -0.39) | 5.33 × 10<sup>-7</sup> | 5.8% | -0.14 (-0.57, 0.29) | 0.52 | -0.42 (-0.91, 0.06) | 0.089 |
| 3 | 139926692 | rs10935361 | NMNAT3 & CLSTN2<sup>a</sup> | T | 30.7% | -0.32 (-0.43, -0.20) | 1.52 × 10<sup>-7</sup> | 31.0% | -0.05 (-0.27, 0.17) | 0.68 | -0.20 (-0.46, 0.06) | 0.13 |
| 7 | 130884634 | rs157916 | LINC-PINT | A | 47.9% | 0.28 (0.17, 0.39) | 4.69 × 10<sup>-7</sup> | 47.1% | -0.05 (-0.24, 0.14) | 0.60 | 0.12 (-0.20, 0.45) | 0.45 |
| 8 | 14866463 | rs76354500 | SGCZ | C | 3.8% | -0.73 (-1.00, -0.46) | 1.54 × 10<sup>-7</sup> | 4.1% | -0.28 (-0.77, 0.20) | 0.26 | -0.55 (-0.98, -0.12) | 0.012 |
| 10 | 79154025 | rs6480922 | ZMIZ1 | T | 20.4% | -0.36 (-0.49, -0.22) | 2.21 × 10<sup>-7</sup> | 20.5% | -0.23 (-0.48, 0.01) | 0.065 | -0.33 (-0.45, -0.22) | 1.42 × 10<sup>-8</sup> |
| 11 | 7194782 | rs11041236 | RBMXL2 & MIR302E<sup>a</sup> | T | 1.2% | -1.28 (-1.78, -0.79) | 4.51 × 10<sup>-7</sup> | 1.9% | 0.52 (-0.20, 1.25) | 0.15 | -0.40 (-2.16, 1.36) | 0.66 |
| Suggestive associations in non-ε4 cases in the discovery series (p<1 × 10<sup>-6</sup>) |  |  |  |  |  |  |  |  |  |  |  |  |
| 11 | 102445120 | rs13377311<sup>b</sup> | TMEM123 | C | 8.0% | 0.73 (0.46, 1.00) | 1.28 × 10<sup>-7</sup> | 5.6% | -0.06 (-0.63, 0.51) | 0.83 | 0.38 (-0.39, 1.15) | 0.34 |

Chr=Chromosome. MA=minor allele; MAF=minor allele frequency; $\beta$ =regression coefficient; CI=confidence interval. For the discovery series and the replication series, $\beta$ values, 95% CIs, and p-values result from linear regression models that were adjusted for age at death, sex, hippocampal region, and the top five principal components of genetic data (the top five principal components were adjusted for only in the discovery series). $\beta$ values are interpreted as the change in mean p-S65-Ub level (on the natural logarithm scale) corresponding to each additional minor allele for the given variant. For the combined analysis, $\beta$ values, 95% CIs, and p-values result from a random effects meta-analysis. <sup>a</sup> The closest genes to the identified variant. <sup>b</sup> Genotyping failed for rs13377311 in the replication series, so the rs10895296 proxy variant was used.

### Combined analysis of discovery-replication series reveals two genome-wide significant hits

In addition to the two-stage GWAS, we conducted a combined analysis of both cohorts as it has been reported to be more efficient than focusing on separate discovery-replication phases.^13^ Using this approach, genome-wide significant associations with p-S65-Ub level were observed for both *APOE* rs429358 (*β*: 0.50, 95% CI: 0.41 to 0.60; *p*=8.67x10^-25^; **Fig. 2B, 3A**), and *ZMIZ1* rs6480922 (*β*: -0.33, 95% CI: -0.45 to -0.22; *p*=1.42x10^-8^; **Fig. 2C, 3A**). We next investigated the potential dependence and interaction between the two genome-wide significant hits in the combined series. In linear regression analysis adjusting for both variants (as well as age at death, sex, and series), relatively consistent associations with p-S65-Ub level were noted for *APOE* rs429358 (*β*: 0.49, 95% CI: 0.39 to 0.59; *p*=5.97x10^-22^) and *ZMIZ1* rs6480922 (*β*: -0.28, 95% CI: -0.40 to -0.16; *p*=2.25x10^-6^). When adding an interaction term between these two variants into the aforementioned linear regression model, some evidence of an interaction was observed (*p*=0.047). Specifically, the association between p-S65-Ub and *ZMIZ1* rs6480922 was stronger for LBD cases without an *APOE4* allele (*β*: -0.38, 95% CI: -0.54 to -0.22; *p*=2.87x10^-6^) compared to cases who did carry *APOE4* (*β*: -0.19, 95% CI: -0.36 to -0.02; *p*=0.029).

### Variants in *APOE* and *ZMIZ1* associate with changes in brain weight and neuropathological burden

Next, we aimed to examine the mechanism by which the identified *APOE* and *ZMIZ1* variants might influence hippocampal p-S65-Ub levels. In this context, it is noteworthy to clarify that the *APOE4* genotype was associated with greater p-S65-Ub levels, while the *ZMIZ1* variant was associated with lesser p-S65-Ub levels (**Fig. 3A**). Both effects were gene-dosage dependent. From earlier studies we know that p-S65-Ub level strongly correlates with the neuropathological burden in LBD and in AD.^8–10^ We thus collected additional quantitative neuropathological measures for all cases (**Supplemental Table 2**) and studied their respective association with the identified variants in the combined series after adjustment for age at death and sex (**Supplemental Table 4**, **Fig. 3B**). It is well established that *APOE4* increases risk for AD and DLB and is known to exacerbate neuropathological burden.^18^ Indeed, brain weight was negatively associated with increased *APOE4* allele count (*p*=6.47x10^-7^). In line, *APOE4* was also associated with an increase of each neuropathological load as measured by αSyn burden (NACP immunostaining) (*p*=3.59x10^-5^), and densities of SPs and NFTs (*p*=2.31x10^-41^ and *p*=7.36x10^-23^) in the hippocampus. This suggests that *APOE4* may cause mitophagy defects, at least in part, through aggravating the neuropathologies. In contrast, the *ZMIZ1* variant was significantly associated with increased brain weight (*p*=0.0006) as well as reduced neuropathological burden of αSyn (*p*=0.031), SP (*p*=1.41x10^-5^), and NFT (*p*=0.001).

**Figure 3.**
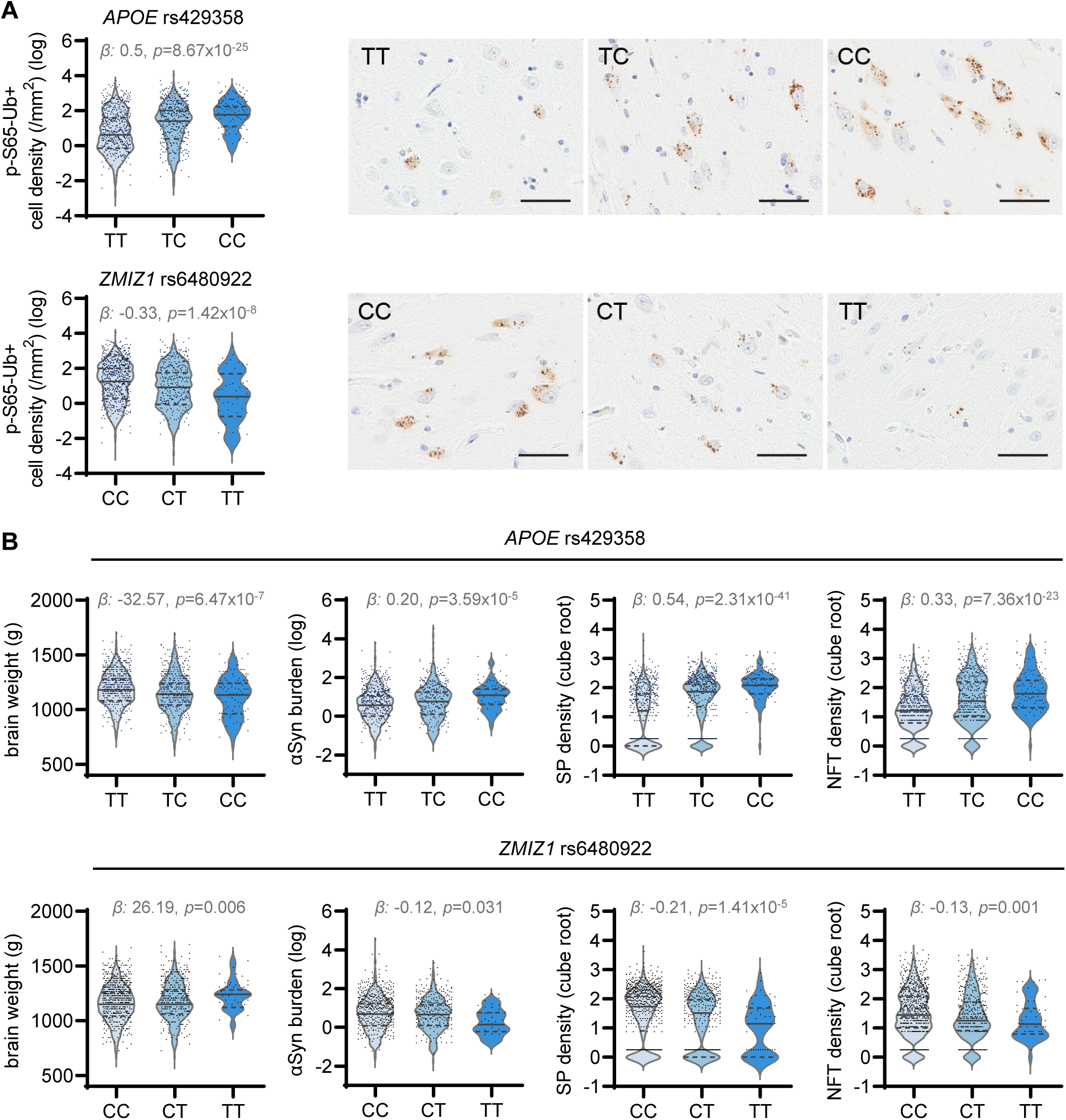
Significant association of *APOE* and *ZMIZ1* variants with p-S65-Ub levels, brain weight, and neuropathological measures in LBD autopsy brain. (**A**) Violin plot of pS65-Ub level (natural logarithm scale) according to number of minor alleles of *APOE* rs429358 and *ZMIZ1* rs6480922 in the combined (discovery and replication) series, together with the corresponding representative images of p-S65-Ub staining in the hippocampus in LBD cases. Scale bar: 50 µm. (**B**) Violin plot of brain weight, αSyn burden (by NACP staining) (natural logarithm scale) and densities of SPs and NFTs (count per microscope field) (cube root scale) according to number of minor alleles of *APOE* rs429358 and *ZMIZ1* rs6480922 in the combined (discovery and replication) series. *β* = regression coefficient beta. SP = senile plaque, NFT = neurofibrillary tangle.

### *APOE4*-associated p-S65-Ub increase in mouse model and iPSC-derived astrocytes independent from neuropathology

To test potential neuropathology-independent *APOE4* effects on mitophagy, we assessed p-S65-Ub levels in experimental animals and cell models using a previously established sensitive MSD electrochemiluminescence assay^15^. First we analyzed brains of Apoe-TR mice, in which the murine *Apoe* gene locus is replaced with human *APOE3* or *APOE4* gene^14^. We detected significantly elevated p-S65-Ub levels in brain lysates of *APOE4* mice at younger age (3 months) when compared to age- and sex-matched *APOE3* mice (**Fig. 4A-B**). Additionally, p-S65-Ub levels were increased in aged *APOE3* mice relative to their younger counterparts but no further escalation in aged *APOE4* mice in comparison to both young *APOE4* and aged *APOE3* mice. As *APOE* is mainly expressed in astrocytes within the central nervous system, we next examined p-S65-Ub levels at baseline and upon mitochondrial stress in human iPSC-derived astrocytes from *APOE3* or *APOE4* homozygous individuals (**Fig. 4C**). Treatment with the mitochondrial stressor CCCP induced significantly higher p-S65-Ub levels in *APOE4* astrocytes compared to *APOE3*, while levels of Ub kinase PINK1 were similar between two genotypes (**Fig. 4D-E**). Altogether, we were able to confirm *APOE4*-associated p-S65-Ub increase in mouse brain and human iPSC models that was independent of disease-related pathology.

**Figure 4.**
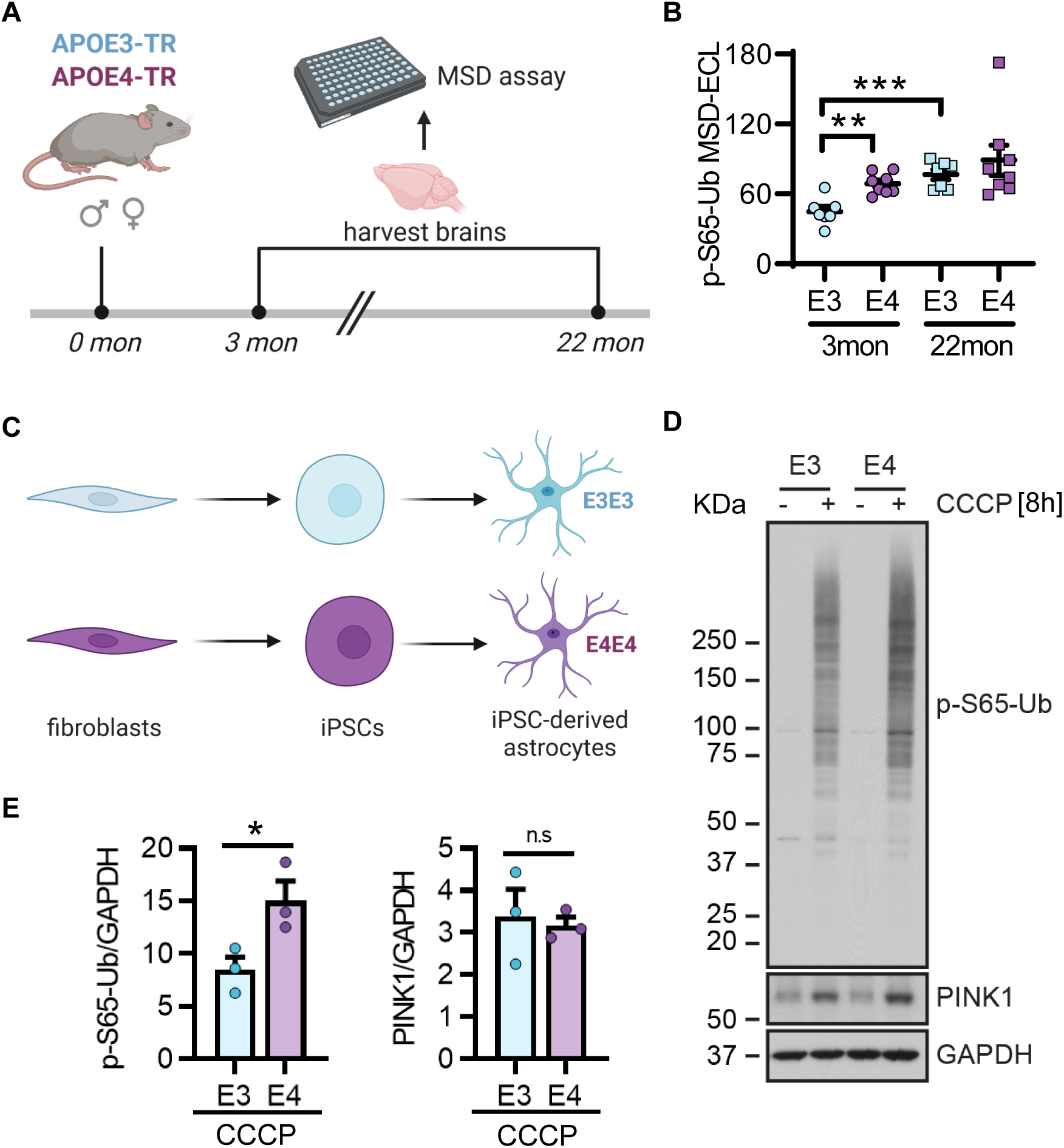
Functional validation of APOE4 effect on mitophagy changes in mouse and iPSC models. **(A)** Brain lysates from 3- and 22-month-old Apoe-TR mice are used to measure p-S65-Ub levels. **(B)** MSD shows significantly increased p-S65-Ub levels in brain lysates from 3-months-old *APOE4* mice compared to age-matched *APOE3* mice. An age-dependent increase of p-S65-Ub levels is also observed in *APOE3* mice. (**C**) Dermal fibroblasts collected from individuals carrying homozygous *APOE3* or *APOE4* are converted into iPSCs, which are then differentiated into astrocytes. (**D**) Representative western blot images of human iPSC-derived astrocytes. (**E**) Western blot results showed that human iPSC-derived homozygous *APOE4* astrocytes have significantly increased p-S65-Ub levels but similar PINK1 levels compared to homozygous *APOE3* astrocytes upon mitochondrial depolarization (8h CCCP treatment). Panel (A) and (C) are created with BioRender.com.

## Discussion

Our GWAS of neuropathologically diagnosed LBD cases uncovered common genetic variants that modify PINK1-PRKN-directed mitophagy. In contrast to binary case-control studies, we here determined levels of the ‘mitophagy tag’ p-S65-Ub, as a novel biological trait. p-S65-Ub serves as a selective and sensitive marker of mitochondrial damage that was exploited as quantitative endophenotype. Our GWAS identified a strong genome-wide significant association with p-S65-Ub on chromosome 19 for rs429358 (*APOE4*), and the combined analysis of discovery and replication cohorts nominated another significant association on chromosome 10, rs6480922, in the *ZMIZ1* gene. Both variants are associated with p-S65-Ub levels in an allele dose-dependent manner, but in different directions: i.e., *APOE* rs429358 is associated with higher, while *ZMIZ1* rs6480922 is associated with lower-than-normal levels of p-S65-Ub. These opposite effects were also reflected in differential associations of both variants with brain weight as well as hippocampal burden of αSyn, Aβ, and p-tau pathologies. This may suggest that p-S65-Ub has greater sensitivity than each neuropathological parameter alone which could allow for not only the identification of genetic modifiers of LBD, but perhaps also resilience to neuropathology.

The *APOE4* allele is the strongest common genetic risk factor for both AD and DLB.^3,19^ A role for *APOE4* in driving Aβ and p-tau deposition is well established^18^, and it has been suggested that *APOE4* is also associated with increased severity and spread of LBs, independently of AD pathology.^20^ p-S65-Ub significantly increases in human brain with age and independently with αSyn and early tau pathology.^8,9,10^ As such it is conceivable that at least some of the *APOE4*-dependent increase in p-S65-Ub could stem from an exacerbated neuropathology, especially as additive and synergistic effects of αSyn, Aβ, and p-tau are emerging from large-scale clinicopathological correlations (Hou X, unpublished). However, due to the very strong association of *APOE4* with both p-S65-Ub and the different neuropathologies in LBD, we unfortunately cannot simply adjust for such neuropathological measures in our statistical models in order to assess whether *APOE4* is associated with p-S65-Ub levels independently of the neuropathology. Therefore, we subsequently undertook an experimental model approach to better elucidate a possible independent association between *APOE4* and p-S65-Ub.

APOE, a major lipid carrier that is predominantly expressed in astrocytes in the brain, broadly impacts lipid homeostasis and thereby affect functions of both mitochondria and lysosomes.^21–23^ As such it is plausible that p-S65-Ub levels are increased through APOE4-dependent changes in lipid metabolism which could lead to increased build-up or decreased clearance of p-S65-Ub labeled mitochondria. To test such biological effect, we utilized Apoe-TR mice that express human *APOE3* or *APOE4* and display the characteristic lipid dysregulation, but no disease related pathology.^14^ p-S65-Ub levels were indeed significantly higher already in brain from young *APOE4* mice (3 months) compared to age-matched *APOE3* mice reaching levels typically seen only at older ages (22 months). This is consistent with our prior work that demonstrated an age-dependent increase of p-S65-Ub in human^8,17^ and recently also in mouse brain (Watzlawik JO, in revision). In addition, analyses of iPSC-derived astrocytes treated with the mitochondrial depolarizer CCCP, which maximally activates PINK1-PRKN signaling, further confirmed higher p-S65-Ub levels in *APOE4* compared to *APOE3* astrocytes. Of note, the Ub kinase PINK1 was similarly stabilized following CCCP treatment, potentially pointing towards a less effective clearance of p-S65-Ub labeled mitochondria in *APOE4* astrocytes. In summary, we found significantly elevated p-S65-Ub levels in complementary experimental models in the absence of αSyn, Aβ, or p-tau deposits indicating additional, pathology-independent effects. Further investigations of the underlying mechanism(s) will be important.

In addition to the *APOE4* genotype, we detected another genome-wide significant variant (rs6480922) located in an intron of the *ZMIZ1* gene that encodes a transcriptional coactivator involved in various signaling pathways.^24^ In humans, pathogenic *ZMIZ1* mutations have been linked to a syndromic neural disorder with intellectual disability and neurodevelopmental delay.^25^ Furthermore, variants in the *ZMIZ1* gene have been identified as risk alleles for various human cancers, autoimmune diseases, and chronic inflammation.^24^ A role of ZMIZ1 for mitophagy has not been described, however, intriguingly, inactivation of the zebrafish ortholog leads to reduced expression of autophagy genes and an accumulation of mitochondrial DNA, which could be indicative of mitophagy defects.^26^ Although we see no evidence of rs6480922 acting as an expression quantitative trait loci status for *ZMIZ1* or neighboring genes, further studies are warranted and should consider cell type specific effects.^27^ Of note, and in contrast to *APOE4*, *ZMIZ1* rs6480922 was associated with lower p-S65-Ub levels, increased brain weight, and reduced neuropathological burden. Follow-up studies are required to test the potential function of ZMIZ1 as a resilience factor to mitochondrial damage and LBD neuropathology.

Our study also has certain limitations. First, the analysis only included LBD cases of Caucasian ancestry, and therefore it will be important for future work to assess genetic risk factors for p-S65-Ub in other ethnic groups. Second, despite a relatively large sample size for a study of neuropathologically-confirmed LBD cases, the possibility of a type II error (i.e., a false-negative finding) is still important to consider, especially for the smaller non-*APOE4* cohort. We cannot conclude that a true association with p-S65-Ub level does not exist simply due to the occurrence of a non-genome-wide significant p-value in our study, and more samples will likely reveal additional significant hits. Third, our proof-of-concept study focused on p-S65-Ub levels in the entire hippocampus. Limiting the analysis to certain vulnerable hippocampal subfields or expansion to different brain regions or even cohorts might shed more light onto shared or distinct neurodegenerative pathways across a spectrum of diseases.

Altogether, capitalizing on a specific and sensitive mitochondrial damage marker, we herein identified *APOE4* and *ZMIZ1* rs6480922 as new mitophagy modifiers in a large series of pathologically confirmed LBD brains. As mitochondrial deficits are early pathological features of LBD, the nominated variants in the current study may ultimately lead to new therapeutic targets that benefit the fast-growing disease population. With the ongoing development of small molecule mitophagy activators, results from this study may help guide future customized therapies for carriers with specific genetic variants. We further demonstrated the versatility of the mitophagy tag as a genetic screening tool and highlighted its relevance and potential as a biomarker of aging and neurodegeneration. Given that p-S65-Ub is also present and detectable in human blood^15^, studies interrogating its suitability to monitor mitochondrial health status in clinical samples are warranted and may help improve diagnosis and prognosis in LBD.

## Data availability

Access to the datasets used and/or analyzed during the current study are available from the corresponding author Dr. Wolfdieter Springer on request.

## Acknowledgements

We are grateful to the patients and their families who made this study possible. We thank Monica Castanedes-Casey and Virginia R. Phillips from the Neuropathology Laboratory for processing human post-mortem tissues and the excellent technical support.

## Funding

WS, OAR, and DWD are members of the American Parkinson Disease Association (APDA) Center for Advanced Research at Mayo Clinic Florida and are further supported by the National Institute of Neurological Disorders and Stroke (NIH/NINDS) Lewy Body Dementia Center Without Walls [U54 NS110435]. WS is additionally supported by NIH [R01 NS085070, R01 NS110085, and R56 AG062556], the Department of Defense Congressionally Directed Medical Research Programs (CDMRP) [W81XWH-17-1-0248], the Michael J. Fox Foundation for Parkinson’s Research (MJFF), the Ted Nash Long Life Foundation, Mayo Clinic Foundation, the Center for Biomedical Discovery (CBD), and the Robert and Arlene Kogod Center on Aging. XH is supported by a pilot grant and a developmental project award from the Mayo Clinic Alzheimer Disease Research Center (ADRC, P30 AG062677) and fellowships awarded by the APDA and Alzheimer’s Association [AARF-22-973152]. FCF is the recipient of fellowships from the Younkin Scholar Program and the APDA and is supported in part by the Florida Department of Health - Ed and Ethel Moore Alzheimer’s Disease Research Program [22A07], the MJFF, a Gerstner Family Career Development Award from the Center for Individualized Medicine (CIM) and an auxiliary award from the CBD at Mayo Clinic. SK is partially supported by the Florida Department of Health - Ed and Ethel Moore Alzheimer’s Disease Research Program [22A05] and a developmental project award from the Mayo Clinic Alzheimer Disease Research Center (ADRC, P30 AG062677). OAR is supported in part by NIH [P50 NS072187, R01 NS078086, and U54 NS100693], Department of Defense [W81XWH-17-1-0249], the ABF, the MJFF, Ted Turner and Family, the Little Family Foundation, and the CIM at Mayo Clinic. DWD is further supported by the Mangurian Foundation Lewy Body Dementia Program at Mayo Clinic. NZ is supported by NIH grants [U19 AG069701 and RF1 AG046205].

## Competing interests

Mayo Clinic, FCF, and WS have filed a patent related to PRKN activators. All other authors declare they have no competing interests. This research was conducted in compliance with Mayo Clinic conflict of interest policies.

## Notes

### Funding Statement

This study was funded by National Institute of Neurological Disorders and Stroke (NIH/NINDS) Lewy Body Dementia Center Without Walls [U54 NS110435] and [RF1 NS085070] as well as a developmental project award from the Mayo Clinic Alzheimer Disease Research Center [ADRC, P30 AG062677] and a fellowship awarded by Alzheimers Association [AARF-22-973152].

### Author Declarations

Ethics committee of Mayo Clinic exempted current study from human subjects regulations given it used only de-identified postmortem brain tissue.

