## Supplemental Materials for "Genome-wide association study identifies *APOE* and *ZMIZ1* variants as mitophagy modifiers in Lewy body disease"

### Supplemental material

**Supplemental Table 1. Subject characteristics**

| | Discovery series (*n*=754) | | Replication series (*n*=258) | |
|---|---|---|---|---|
| **Variable** | ***n*** | **Median (minimum, maximum) or No. (%) of cases** | ***n*** | **Median (minimum, maximum) or No. (%) of cases** |
| **Age at death (years)** | 754 | 78 (48, 99) | 258 | 76 (56, 95) |
| **Sex (Male)** | 754 | 450 (59.7%) | 258 | 186 (72.1%) |
| **LBD subtype** | 754 | | 258 | |
| Brainstem | | 108 (14.3%) | | 0 (0.0%) |
| Transitional | | 263 (34.9%) | | 91 (35.3%) |
| Diffuse | | 383 (50.8%) | | 167 (64.7%) |
| **Braak (tau) stage** | 754 | | 205 | |
| 0 | | 16 (2.1%) | | 6 (2.9%) |
| I | | 32 (4.2%) | | 10 (4.9%) |
| II | | 132 (17.5%) | | 43 (21.0%) |
| III | | 199 (26.4%) | | 51 (24.9%) |
| IV | | 117 (15.5%) | | 71 (34.6%) |
| V | | 109 (14.5%) | | 14 (6.8%) |
| VI | | 149 (19.8%) | | 10 (4.9%) |
| **Thal phase** | 754 | | 258 | |
| 0 | | 101 (13.4%) | | 42 (16.3%) |
| 1 | | 65 (8.6%) | | 23 (8.9%) |
| 2 | | 41 (5.4%) | | 23 (8.9%) |
| 3 | | 151 (20.0%) | | 68 (26.4%) |
| 4 | | 77 (10.2%) | | 33 (12.8%) |
| 5 | | 319 (42.3%) | | 69 (26.7%) |
| **p-S65-Ub level** | 754 | 3.23 (0.06, 38.02) | 258 | 2.33 (0.11, 25.25) |

**Supplemental Table 2. Summary of additional neuropathological characteristics**

| Variable | *n* | Median (minimum, maximum) |
|---|---|---|
| Brain weight (g) | 1005 | 1160 (660, 1630) |
| αSyn burden | 717 | 1.95 (0.18, 63.84) |
| SP density (count per microscope field) | 1004 | 4.50 (0.00, 38.33) |
| NFT density (count per microscope field) | 1009 | 2.75 (0.00, 40.50) |

**Supplemental Table 3a. Genotype counts and frequencies in the discovery series for the 8 variants displaying genome-wide significant and suggestive associations**

| Variant | Minor allele count and frequency | Major allele count and frequency | Genotype 1 count and frequency | Genotype 2 count and frequency | Genotype 3 count and frequency |
|---|---|---|---|---|---|
| rs429358 | C: 452 (30.0%) | T: 1032 (70.0%) | TT: 373 (49.5%) | TC: 310 (41.1%) | CC: 71 (9.4%) |
| rs6712544 | A: 73 (4.8%) | C: 1435 (95.2%) | CC: 683 (90.6%) | CA: 69 (9.2%) | AA: 2 (0.3%) |
| rs10935361 | T: 463 (30.7%) | C: 1045 (69.3%) | CC: 358 (47.5%) | CT: 329 (43.6%) | TT: 67 (8.9%) |
| rs157916 | G: 723 (47.9%) | A: 785 (52.1%) | AA: 203 (26.9%) | AG: 379 (50.3%) | GG: 172 (22.8%) |
| rs76354500 | C: 57 (3.8%) | T: 1451 (96.2%) | TT: 700 (92.8%) | TC: 51 (6.8%) | CC: 3 (0.4%) |
| rs6480922 | T: 308 (20.4%) | C: 1200 (79.6%) | CC: 474 (62.9%) | CT: 252 (33.4%) | TT: 28 (3.7%) |
| rs11041236 | T: 18 (1.2%) | C: 1490 (98.8%) | CC: 736 (97.6%) | CT: 18 (2.4%) | TT: 0 (0.0%) |
| rs13377311 (non-ε4 cohort) | T: 60 (8.0%) | C: 686 (92.0%) | CC: 316 (84.7%) | CT: 54 (14.5%) | TT: 3 (0.8%) |

**Supplemental Table 3b. Genotype counts and frequencies in the replication series for the 8 variants selected for inclusion in the replication series**

| Variant | Minor allele count and frequency | Major allele count and frequency | Genotype 1 count and frequency | Genotype 2 count and frequency | Genotype 3 count and frequency |
|---|---|---|---|---|---|
| rs429358 | C: 149 (29.1%) | T: 363 (70.9%) | TT: 134 (52.3%) | TC: 95 (37.1%) | CC: 27 (10.5%) |
| rs6712544 | A: 30 (5.8%) | C: 486 (94.2%) | CC: 229 (88.8%) | CA: 28 (10.9%) | AA: 1 (0.4%) |
| rs10935361 | T: 160 (31.0%) | C: 356 (69.0%) | CC: 120 (46.5%) | CT: 116 (45.0%) | TT: 22 (8.5%) |
| rs157916 | G: 243 (47.1%) | A: 273 (52.9%) | AA: 64 (24.8%) | AG: 115 (44.6%) | GG: 79 (30.6%) |
| rs76354500 | C: 21 (4.1%) | T: 491 (95.9%) | TT: 236 (92.2%) | TC: 19 (7.4%) | CC: 1 (0.4%) |
| rs6480922 | T: 106 (20.5%) | C: 410 (79.5%) | CC: 161 (62.4%) | CT: 88 (34.1%) | TT: 9 (3.5%) |
| rs11041236 | T: 10 (1.9%) | C: 506 (98.1%) | CC: 248 (96.1%) | CT: 10 (3.9%) | TT: 0 (0.0%) |
| rs10895296 (non-ε4 cohort) | G: 15 (5.6%) | A: 253 (94.4%) | AA: 119 (88.8%) | AG: 15 (11.2%) | GG: 0 (0.0%) |

[a] Genotyping failed for rs13377311 in the replication series, so the rs10895296 proxy variant was used.

**Supplemental Table 4. Associations of *APOE* rs429358 and *ZMIZ1* rs6480922 with additional neuropathological characteristics**

| | | Association with *APOE* rs429358 | | Association with *ZMIZ1* rs6480922 | |
|---|---|---|---|---|---|
| **Variable** | ***n*** | ***β* (95% CI)** | **p-value** | ***β* (95% CI)** | **p-value** |
| Brain weight (g) | 1005 | -32.57 (-45.34, -19.81) | $6.47 \times 10^{-7}$ | 26.19 (11.35, 41.03) | 0.0006 |
| αSyn intensity | 717 | 0.20 (0.11, 0.30) | $3.59 \times 10^{-5}$ | -0.12 (-0.22, -0.01) | 0.031 |
| SP density | 1004 | 0.54 (0.47, 0.62) | $2.31 \times 10^{-41}$ | -0.21 (-0.30, -0.12) | $1.41 \times 10^{-5}$ |
| NFT density | 1009 | 0.33 (0.27, 0.39) | $7.36 \times 10^{-23}$ | -0.13 (-0.21, -0.05) | 0.001 |

β=regression coefficient; CI=confidence interval. β values, 95% CIs, and p-values result from linear regression models that were adjusted for age at death and sex. β values are interpreted as the change in the mean neuropathological variable (on the natural logarithm scale for αSyn burden and on the cube root scale for SP and NFT density) corresponding to each additional minor allele for the given variant.

### Supplemental Methods

#### Immunohistochemistry of human brain tissue

The tissue blocks used in the study were collected and prepared in a very uniform way ensuring consistent and comparable hippocampal region in all samples. The paraffin embedded brain tissue was cut into 5 micron sections and allowed to dry overnight at 60°C. Slides were deparaffinized and rehydrated, followed by antigen retrieval in steaming deionized water. After blocking with 0.03% hydrogen peroxide and 5% normal goat serum (Invitrogen, 16210072), sections were incubated with primary antibodies against p-S65-Ub (in-house, 1:650), followed by rabbit-labeled polymer HRP (Agilent, K4011) at room temperature. Peroxidase labeling was visualized with the DAB chromogen before sections were counterstained and coverslipped.

#### Meso Scale Discovery electrochemiluminescence assays of mouse brain tissue

96-well plates were coated with the p-S65-Ub antibody (Cell Signaling Technology, 62802, 1:100) in carbonate coating buffer (200 mM Na2CO3, pH9.6), incubated overnight at 4°C, and washed before use. Following 1 h blocking with 1% BSA in TBST, 30 µg of brain lysates were diluted in 1% BSA in TBST to 30 µl and loaded into each well in duplicate. Samples were incubated for 2 h in the plate at room temperature while shaking. Plates were then washed again before incubating with the total Ub detection antibody (Thermo Fisher, 14-6078-82, 1:100) for 2 h at room temperature. After 3 washes, SULFO TAG detection antibody (Meso Scale Discovery, R32AC, 1:500) was incubated in the plate for 1 h at room temperature. After the final wash, MSD Gold read buffer (Meso Scale Discovery, R92TG) was added to each well, followed by plate reading on the MESO QuickPlex SQ 120 (Meso Scale Diagnostics, Rockville, MD, USA)

#### Differentiation of human iPSCs into astrocytes

To generate neural progenitor cells (NPCs), the iPSCs clumps were cultured in neural induction medium (Stemcell Technologies) in suspension for 5-7 days to initiate neurosphere formation. Next, neurospheres were seeded onto Matrigel-coated dishes and cultured in neural induction medium for another 5-7 days to induce neural rosette formation. Neural rosettes were isolated as a single cell suspension and re-plated onto Matrigel-coated dishes in neural induction medium. The medium was then replaced to neural progenitor cell medium (Stemcell Technologies) and cultured for additional 10-14 days to induce NPCs differentiation. For astrocyte differentiation, NPCs were cultured on PDL-coated plates in astrocyte differentiation medium composed of astrocyte medium (ScienCell) with CNTF (10 ng/ml), BMP4 (10 ng/ml) and Heregulin-β (10 ng/ml) (all from Stemcell Technologies). At day 30, the NPC-derived astrocytes were treated and then harvested for western blot analyses.

#### Western blot of iPSC-derived astrocytes

Cells were lysed in RIPA buffer containing protease inhibitor cocktail and phosphatase inhibitors (Sigma-Aldrich, 11697498001 and 04906837001). Cell lysates were incubated on ice for 30 min and centrifuged in cold for 15 min at 14,000 rmp to remove the cell debris. Supernatant were collected and protein concentrations were determined by BCA assay (Thermo Fisher, 23225). Cell lysates containing 20 µg of protein were diluted in Laemmli buffer (62·5 mM Tris, pH 6·8, 1·5% SDS, 8·33% glycerol, 1·5% β-Mercaptoethanol, 0·005% bromophenol blue) and boiled at 95°C for 5 min. The denatured samples were then run on Tris-Glycine gels (Invitrogen, EC60485BOX). After transferring protein onto PVDF membranes (Millipore Sigma, IPVH00010), membranes were blocked in 5% skim milk (Genesee, 20-241) and incubated with primary antibodies against p-S65-Ub (in-house, 1:10,000), PINK1 (Cell Signaling Technology, 6946; 1:1,000), and GAPDH (Meridian Life science, H86504M; 1:150,000) overnight at 4°C, followed by secondary HRP-conjugated antibodies (Jackson ImmunoResearch, 711-035-152 and 715-035-150, 1:10,000) for 1 h at room temperature. Protein bands were visualized using Immobilon Western Chemiluminescent HRP Substrate (Millipore Sigma, WBKLS0500) and Blue Devil Lite X-ray films (Genesee Scientific, 30-810L).

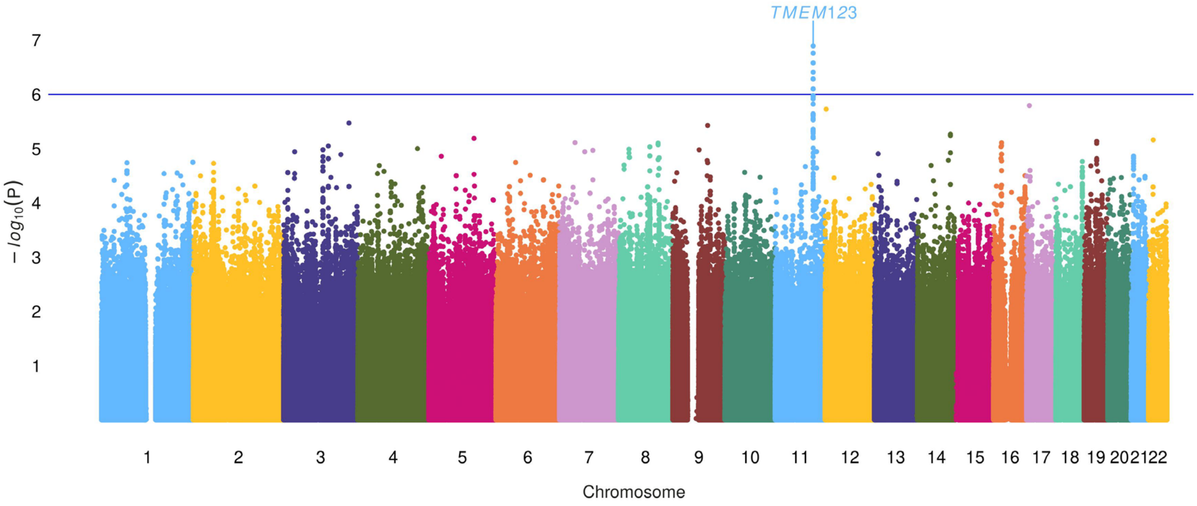

**Supplemental Figure 1. GWAS analysis in LBD autopsy brain from non-*APOE4* carriers.** Manhattan plot showing genome-wide p-values of association with p-S65-Ub levels in non-*APOE4* carriers. The Y axis shows -log10 p-values of 8,696,291 SNPs and the X axis shows their chromosomal positions. The blue solid line indicates the p-value threshold used to define a "suggestive" association ($p<1x10^{-6}$).
